# CADA: Phenotype-driven gene prioritization based on a case-enriched knowledge graph

**DOI:** 10.1101/2021.03.01.21251705

**Authors:** Chengyao Peng, Simon Dieck, Alexander Schmid, Ashar Ahmad, Alexej Knaus, Maren Wenzel, Laura Mehnert, Birgit Zirn, Tobias Haack, Stephan Ossowski, Matias Wagner, Teresa Brunet, Nadja Ehmke, Magdalena Danyel, Stanislav Rosnev, Tom Kamphans, Guy Nadav, Nicole Fleischer, Holger Fröhlich, Peter Krawitz

**Author notes:** ^*^Correspondence.

## Abstract

Many rare syndromes can be well described and delineated from other disorders by a combination of characteristic symptoms. These phenotypic features are best documented with terms of the human phenotype ontology (HPO), which is increasingly used in electronic health records (EHRs), too. Many algorithms that perform HPO-based gene prioritization have also been developed, however, the performance of many such tools suffers from an overrepresentation of atypical cases in the medical literature. This is certainly the case if the algorithm cannot handle features that occur with reduced frequency in a disorder. With CADA we built a knowledge-graph that is based on case annotations and disorder annotations and show that CADA exhibits superior performance particularly for patients that present with the pathognomonic findings of a disease. Crucial in the design of our approach is the use of the growing amount of phenotypic information that diagnostic labs deposit in databases such as ClinVar. By this means CADA is an ideal reference tool for differential diagnostics in rare disorders that can also be updated regularly.

## 1 Introduction

Deep phenotyping of patients with suspected rare genetic disorders by HPO terminology has become the de facto standard and is the prerequisite for several algorithms that prioritize potential disease genes [1, 2, 3, 4]. Since most of the current approaches are still heavily based on disease annotations and not case annotations, many of these tools have become a victim of their own success if they do not take into consideration how frequently a clinical feature occurs: An entry in OMIM evolves over time and accumulates also clinical features that occur rarely: A novel disease-gene-association for a monogenic disorder usually requires three or more unrelated patients with a similar phenotype and mutations in the same gene for a publication in a peer-reviewed journal. After this initial report, often a follow-up study is published a few months or years later that delineates additional clinical features of patients with disease-causing mutation in the same gene. Ideally, such a paper distinguishes between cardinal symptoms of the disorder and those that occur less frequently. Additional case reports are usually just published for patients with an atypical presentation, while most characteristic cases will rather be submitted to databases such as ClinVar [5].

In early algorithms for semantic similarity searches, such as the phenomizer, the specificity of a term is reflected by its information content (IC), which is defined as the negative natural logarithm of the frequency a term has been used to annotate a disease [3]. This approach, however, would result in comparable similarity scores for a gene, no matter whether a patient presents with the two pathognomonic findings present in almost all individuals with this disease, or two rarely occurring features of similar IC.

From 2019, the HPO project also adds metadata to disease annotations which includes the frequency of a clinical feature in a person with a specific disease, however, this data, especially on the gene annotation level is still highly incomplete and inconsistent in its methodology. Nevertheless gene prioritization algorithms stand to benefit significantly from this information and should be ready to include it, as it is further improved in the future.

Shen et. al. [6] showed that graph embeddings of HPO worked well for comparing phenotypes. We extend this approach to also include **C**ase **A**nnotations, as well as **D**isease **A**nnotations (CADA). With this we obtain a graph which can be embedded to perform gene prioritization. Compared to previous methods this graph based approach has the advantage of being weightable with frequency information.

## 2 Materials

### 2.1 Clinical cases

We collected 4,714 clinical cases with varied numbers of phenotypic features (terms in Human Phenotype Ontology) and a genetic diagnosis on a single causal gene from our collaborators and Clinvar. Only submissions in Clinvar with pathogenic and likely pathogenic clinical significance were included in the study and submissions with identical submitters, Hpo terms and disease-causing genes were considered as a single case.

### 2.2 Human Phenotype Ontology

The Human Phenotype Ontology (Hpo) provides a standardized and controlled vocabulary of human phenotypic abnormalities. In Hpo, phenotypic terms are arranged in a directed acyclic graph (DAG) and are related to their parent terms by is a relationships. In our study, we used the Human Phenotype Ontology released on 2020-03-27, containing 14,586 human phenotypic terms and 18, 416,0 hierarchical relationships between these terms.

### 2.3 Gene-phenotype annotations

The Hpo team also provides an annotation file that provides links between genes and Hpo terms. This mapping is based on data mining of resources such as OMIM and Orphanet. In detail, 4, 315 disease-causing genes and 169, 281 unique gene-Hpo term associations are included in our study.

Phenopackets, each consisting of a patient-specific list of HPO-terms and a disease gene, were compiled from electronic patient records of 4714 molecularly confirmed cases [7]. Pathogenic mutations and HPO-terms of an increasing number of these cases have also been submitted to ClinVar, which supports phenotype-rich submissions since 2017 [5]. Of these 4714 cases, we obtained 2577 directly from ClinVar and 2137 where contributed by clinical collaborators.

## 3 Methods

### 3.1 Encoding the data

Comparing nominal data is difficult as there is no mathematical basis to predict similarity. For many problems in the past, embedding the data into a vector space has proven as a good way to allow for statistical computation on nominal data. [8] For the purpose to measure similarity between phenotypes and genes, we embedded the nominal data encoded in Hpo and the associated gene for each phenotype. There are several methods of embedding an ontology into a vector space, however it is worth noting that Hpo only utilises one type of edge and therefore can also be read as a simple graph, with edge pairs instead of triples. Shen et.al. [6] showed that this approach worked well for embedding Hpo.

As opposed to Shen et.al. we also add in gene associations and obtain a graph *G* with two types of nodes. *V*_*P*_, the set of phenotypes present in Hpo and *V*_*G*_, the set of disease-causing genes. There are two sets of edges in the graph, phenotype to phenotype edges *E*_*P P*_ ⊆ {(*p*_1_, *p*_2_)|*p*_1_, *p*_2_ ∈ *V*_*P*_} and phenotype to gene edges *E*_*P G*_ ⊆ {(*p, g*)|*p* ∈ *V*_*P*_, *g* ∈ *V*_*G*_}. So the Graph encoding all relationships is *G* = (*V*_*P*_ ∪ *V*_*G*_, *E*_*P P*_ ∪ *E*_*P G*_).

With this definition we are now able to read in a case *C*, which usually consists of a list of phenotypes *P*_*C*_ ⊂ *V*_*P*_ and the diagnosed disease causing gene *g*_*C*_ ∈ *V*_*G*_ as a set of edges *E*_*C*_ = {(*p, g*_*C*_)|*p* ∈ *P*_*C*_}. Easily allowing us to extend our graph *G* by the information present in the case (see Figure 1).

**Figure 1:**
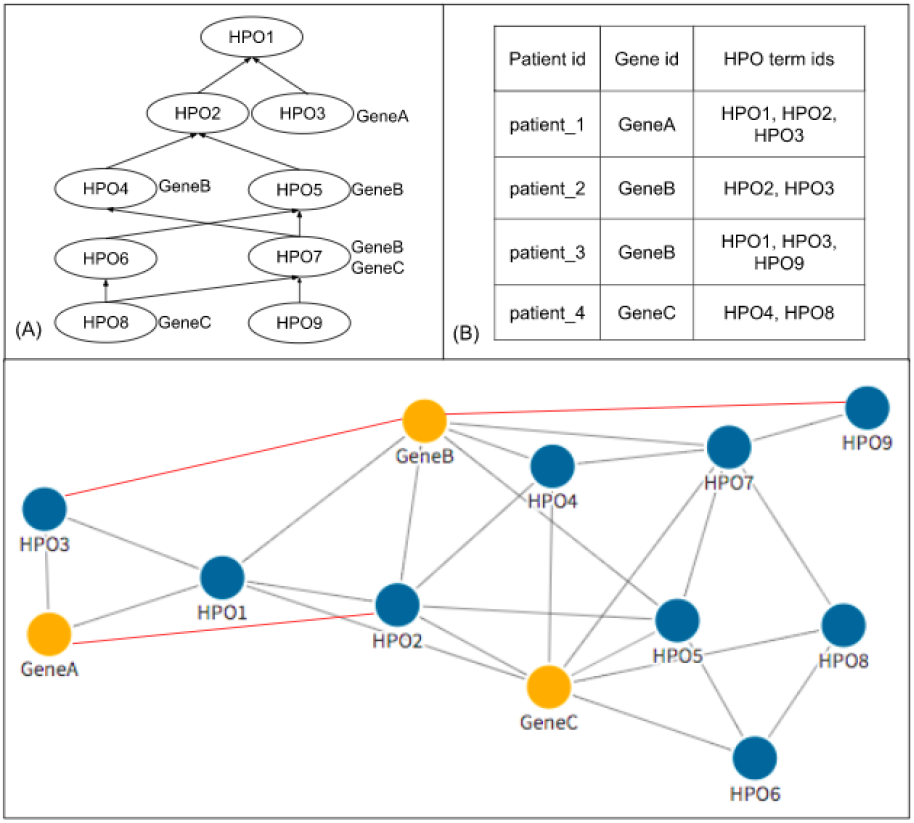
General workflow of encoding the data into the graphs. The initial network *G*_0_ was constructed from the DAG structure and genephenotype annotations from HPO in (A). It consists of the nodes and grey edges in (C). The network was further extended by red edges in by clinical cases in (B).

### 3.2 Embedding the data into a vector space

With the data represented as a graph, *G*_0_, we used Node2Vec to create the vector space embedding [9]. For this purpose, Node2Vec first starts (weighted) random walks on the graph *G*_0_ from each of the nodes. These random walks are interpreted as words that can be embedded into an Euclidean space using a SkipGram neural network, which is an essential part of the Word2Vec method [8]. More specifically, we aim to maximize the probability of a node *v*’s context *R* within a contextual window of length *c*:

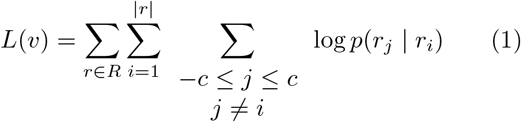

Here *r*_*i*_ denotes the *i*-th word (i.e. node sequence) generated by a random walk. *p*(*r*_*j*_ |*r*_*i*_) is the output of the SkipGram neural network that is defined with a softmax function

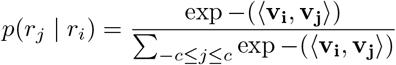

where **v**_**i**_, **v**_**j**_ are vector representations of words *r*_*i*_ and *r*_*j*_ in the hidden layer. Notably, the SkipGram neural network is trained with one-hot vector encoding of word pairs as input. The network aims for learning the probability of observing word *r*_*j*_ in the context (i.e. in the “neighborhood”) of *r*_*i*_ by maximizing ΣΣ_*v*_ *L*(*v*) over all nodes *v* in the graph *G*_0_. We refer to [8] for more details about SkipGram.

To train the Node2Vec model, we split the 4714 patients into a training, validation and test sets with the ratios 60%, 20% and 20%. Note, that *G*_0_ does not contain any case data initially and is only constructed from HPO and the mapping of terms to genes. Cases from the training set were added gradually into *G*_0_. We will denote with e.g. *G*_*p*_ that *p*% of the training patients were added into the graph. The Node2Vec model was trained on *G*_0_, *G*_25_, *G*_75_ and *G*_100_, where hyperparameter optimisation was performed for each of them using the validation set. The Optuna [10] library was used for a Bayesian hyperparameter optimisation. A detailed list of tuned hyperparameters can be found in the Supplemental material.

### 3.3 Using Edge Confidences

In principle each gene to phenotype association should be weighted by clinical case frequency. That means there is a weighting function *h* : *E*_*P G*_ → [0, 1]. Accordingly, for a given node *v* the probability to reach any direct neighbor *q* during a one-step random walk is then

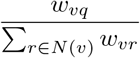

where *N* (*v*) denotes the neigborhood of node *v* and *w*_*vq*_ the weight of the edge *v* →*q*.

Unfortunately, frequencies of gene to phenotype associations are not known on the population level. However, in order to test the principle concept we optionally tested an approach, in which we simply incremented for every gene to phenotype association reported in patients in our training data the weight of an edge by 1.

### 3.4 Link prediction

Node2Vec learns a function *f* : *V*→ ℝ^*d*^ that embeds nodes into a vector space. The problem of disease gene prioritization can be interpreted as as a link prediction task between phenotype and gene nodes. This can be achieved by measuring the similarity of putative disease genes to phenotypes in the vector space via the dot product. Hence, for any new case *C* with a set of phenotypes *P*_*C*_ ⊂*V*_*P*_, a ranking of genes *g* can be achieved via:

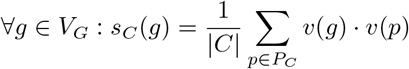

Therefore we can rank genes for each case and compute a top-N accuracy for the test set.

## 4 Results

### 4.2 Robustness of the randomized aspects

As the embedding method is based on a random walk the edge embeddings obtained from Node2Vec will be slightly different each time they are created. Therefore each experiment was repeated with 10 different embeddings obtained from the same graph. Results comparing top-*N* accuracies are average results of these 10 embeddings and have error margins (signified by error bars in figures) associated with them. However the error margins for this method were vanishingly small and around 1% for each top-*N* accuracy. This shows that the method, despite its randomization, is highly robust.

### 4.2 Effects of adding case data

Even without using our weighting scheme, adding the case data into our graph *G*_0_ before the embedding improves top-*N* accuracy significantly for validation cases (Figure 2). The model from unweighted *G*_100_ achieves the best validation results. Similarly, the weighted graph models were also validated through a same approach, among which the weighted *G*_100_ model achieves the best validation results. To test the performance of unweighted and weighted models, we evaluated the *G*_0_, unweighted *G*_100_ and weighted *G*_100_ models with the testing set of 943 patients. Figure 3 shows that all top-N accuracy scores improve around 7-10% by introducing new associations from cases annotations. Moreover, by adding the very simple weighting scheme we propose, the results further improve 3-4%.

**Figure 2:**
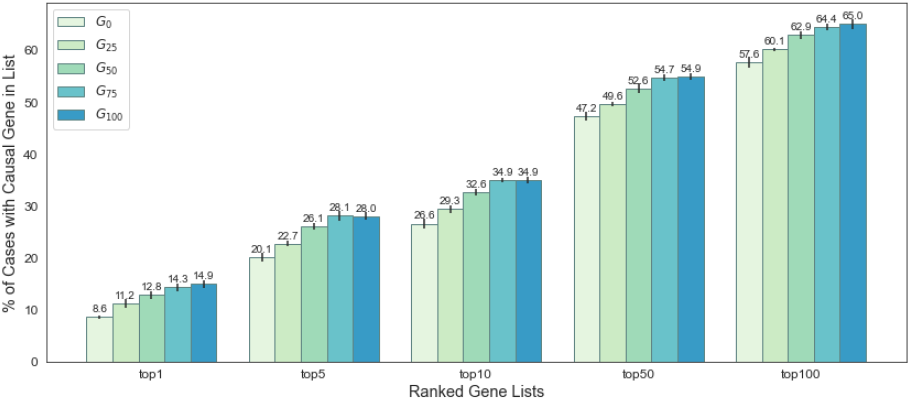
Validation accuracy with standard errors during the graph extension.

**Figure 3:**
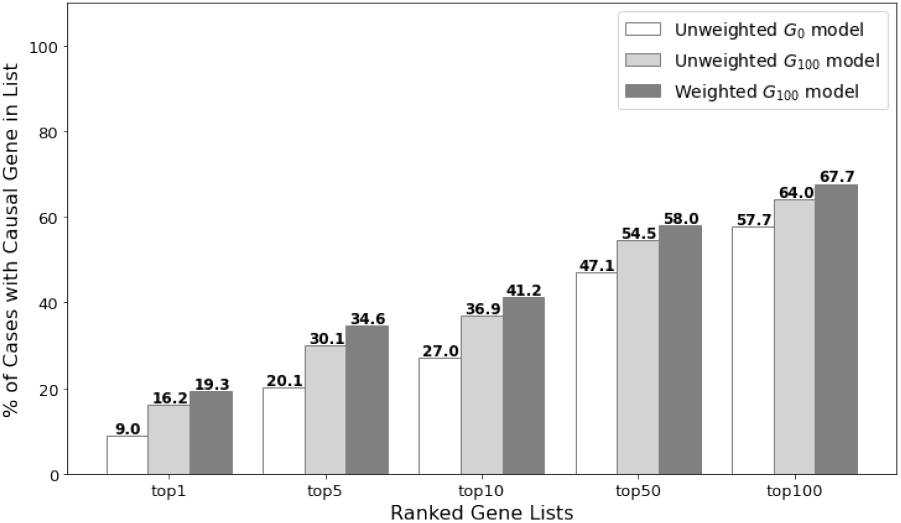
Performance comparison of unweighted and weighted models. The performance of unweighted *G*_0_, *G*_100_ and weighted *G*_100_ models was assessed on 943 testing patients by topN accuracy metrics.

### 4.3 Comparison to other methods

Since the weighting scheme is purely heuristic, we used the unweighted *G*_100_ as the final model to compare with other gene prioritization tools on our test data. However, the restrictions some of the other tools have make a direct comparison difficult. Gado, for instance, can only handle a subset of the phenotypes present in HPO. Thus, it was unable to recognise phenotypic features for around 200 of our test cases. AMELIE requires a pre-selected list of at most 1,000 genes to prioritise, which represents less than one fourth of the 4,315 known disease-causing genes we collected. Only Phen2Gene had directly comparable capabilities to CADA. Therefore, we compared our model to Phen2Gene with the complete list of genes (Figure 4 A) and performed an additional test where 1000 randomly selected genes, including the target gene, where prioritized by CADA, Phen2Gene and AMELIE (Figure 4 B). The comparison tests show that Cada outperforms the other tools even with the unweighted setup.

**Figure 4:**
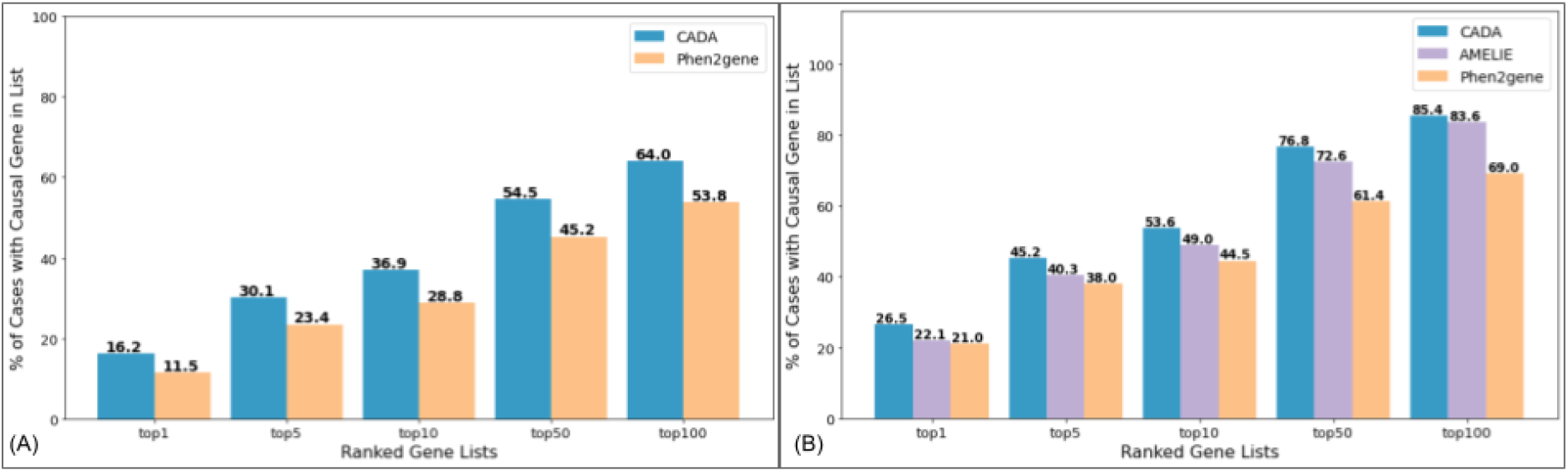
Performance comparison on testing patients. (A) Based on phenotypes alone, CADA was compared with Phen2gene (B) Based on phenotypes and a pre-selected 1,000 gene list, CADA was compared with AMELIE and Phen2gene.

The comparison tests show that Cada outperforms the other tools even with the unweighted setup under both tasks on our test cases. With further improvements when adding in our experimental weighting scheme, the advantage of CADA will be more noticeable. However, Phen2Gene also has further capabilities of identifying potential new disease causing genes. Whilst this wouldn’t affect performance in the experimental setup where a list of 1000 genes was given, it will make the prioritization task naturally harder for Phen2Gene in the general setup.

### 4.4 Performance comparison for gene prevalence groups

To further study how the prevalence of a gene affects its performance by our model, disease-causing genes in case annotations were classified into three prevalence groups based on their frequencies in our case data: common (frequency*>*= 20), rare (5 *<*=frequency*<*= 19), ultra rare (frequency*<*= 4). The number of genes in three prevalence groups and their corresponding patient numbers among training cases are shown in Table 1.

**Table 1:**
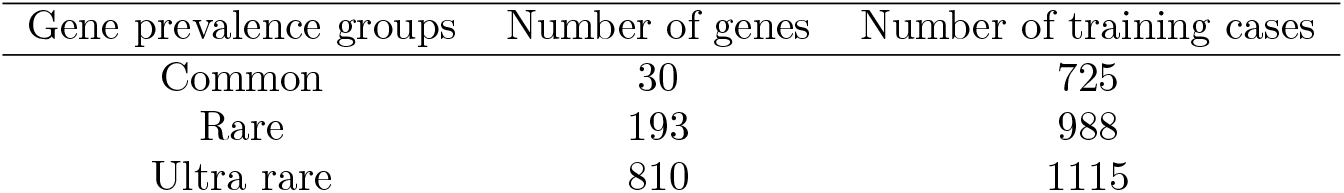
The sum of genes and cases within three prevalence groups.

For the graph extension process, Figure 5A presents the overall distribution of introduced associations among three prevalence groups. Divided by the number of cases and genes in Table 1, the overall distribution was converted to the average distribution of a case (Figure 5 B) and a gene (Figure 5 C) within three prevalence groups.

**Figure 5:**
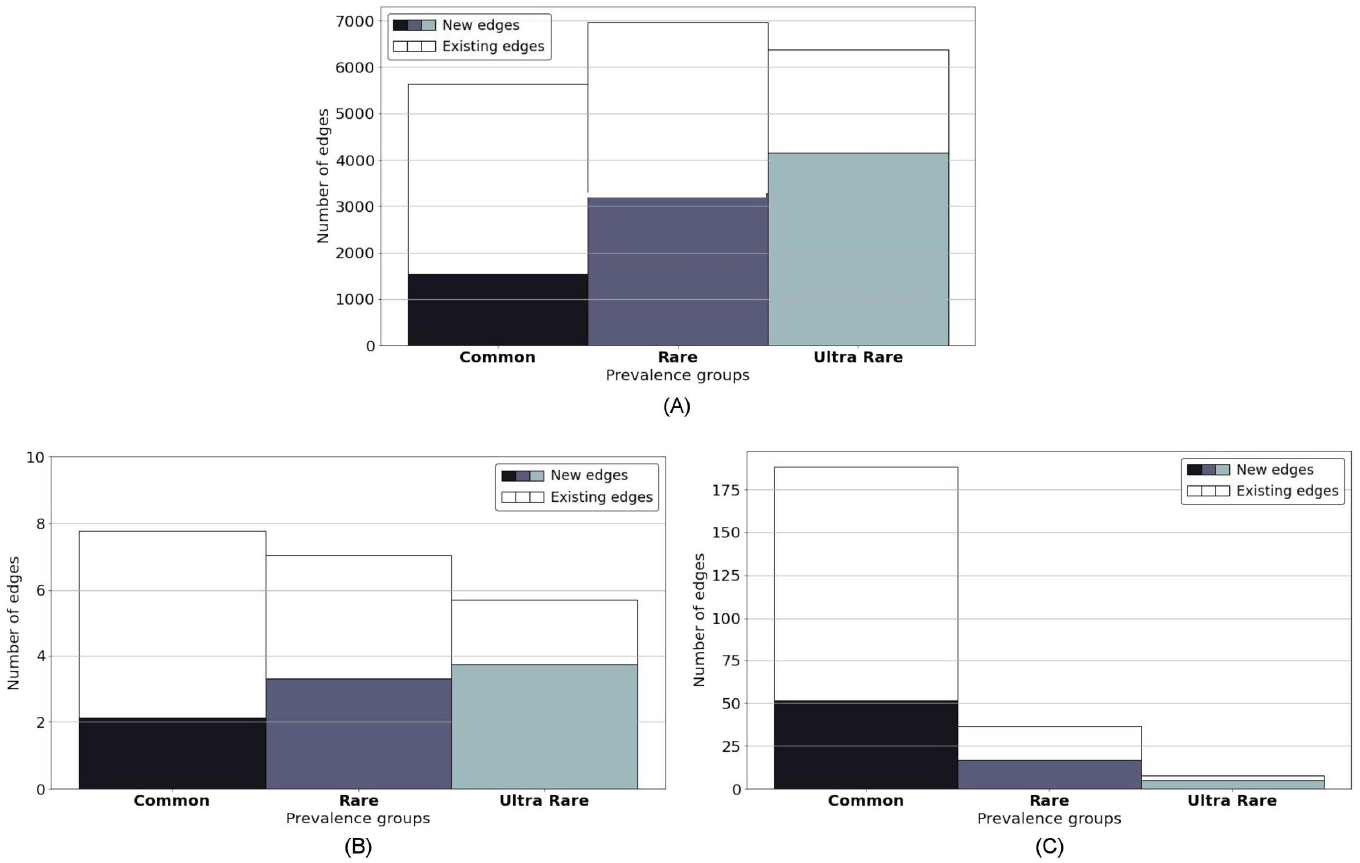
Introduced associations within prevalence groups. (A) The overall distribution of introduced associations within prevalence groups. (B) The average distribution of introduced associations per case. (C) The average distribution of introduced associations per gene.

The performance of test patients was also evaluated accordingly within above-mentioned groups before and after the graph extension. As illustrated in Figure 6, the bars show the accuracy of the *G*_100_ model with white markers indicating the one from the *G*_0_ model. The significant improvement for common genes might result from the forming of abundant new edges on them during the process, as showed in Figure 5 C.

**Figure 6:**
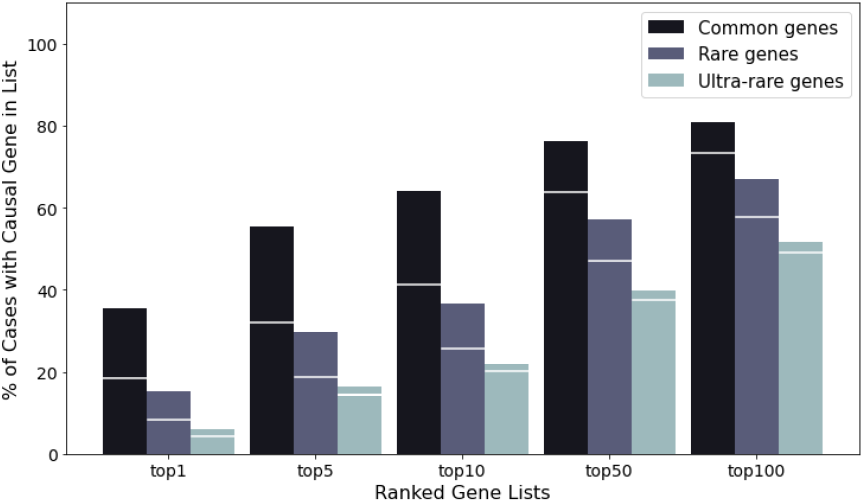
Performance improvement of prevalence groups during the graph extension. The bars show the accuracy of the *G*_100_ model with white markers indicating the accuracy of the *G*_0_ model.

## 5 Discussion

CADA’s underlying graph structure and embedding strategy will enable improvements in the future, also due to its modular design.

Whilst already obtaining comparable results to current tools without weighting the graph, the potential of weighting edges with frequency information is another big advantage of this graph-based approach. Even with a very simple heuristic we were able to improve results significantly. With Hpo currently working on adding frequency information to their database and resources like Orphanet conducting research into frequencies there is high potential for improving this method with a more sophisticated weighting scheme.

Another promising avenue is the rapidly developing field of graph embeddings. Node2Vec was the current most suitable embedding tool we used, however this is a rapidly evolving field, as it has many applications even far beyond medical research. With the current setup for CADA the graph embedding tool can easily be replaced in the future if more promising tools are published.

Robinson, *et al*. recently introduced a framework for estimating posttest probabilities based on likelihood ratios for genotype-phenotype data [11]. By this means, the contribution of each phenotypic feature to a suggested diagnosis can be computed, which is particularly helpful for the clinical interpretation of the results. While LIRICAL is working by default with disease prevalences as pretest probability, it hast also been suggested that other priors e.g. the output of CADA, could be used to refine the output.

In future research we would like to extend the underlying Graph used by CADA with Gene to Gene links to allow for discovery capabilities similarly to Phen2Gene.

The code for CADA, can be found here: https://github.com/Chengyao-Peng/CADA. This code can be used to process a single case in seconds on a regular laptop via commandline, allowing for large scale reprocessing of cases.

Furthermore we’re making this tool available to anyone via a web interface at https://cada.gene-talk.de/webservice/. This version will be updated with new ClinVar cases on a regular basis, and is therefore expected to improve over time.

## Data Availability

All Data used in this study can be found in the corresponding github repository.

https://github.com/Chengyao-Peng/CADA

## 6 Data Availability

The Data used in this paper can be found at https://github.com/Chengyao-Peng/CADA.

